# Kidney patients remain at increased risk for succumbing to COVID-19

**DOI:** 10.1101/2022.06.12.22276220

**Authors:** Rodinde Hendrickx, Mads Jellingsø, Morten OA Sommer

## Abstract

Immunocompromised patients have been at an increased risk of succumbing to COVID-19 already since the beginning of the pandemic. Here we analyzed data from patients with end stage renal disease, including those on dialysis and patients with a kidney transplant, and compared them to the general population. We found that kidney patients remain at increased risk of succumbing to COVID-19 despite all available countermeasures. The analyses underline the need for additional protection for this vulnerable population.

## Main Body

Over the last two years much progress has been made towards protection and treatment against COVID-19. These therapeutic options have dramatically increased survival rates and improved prognosis for many patient groups. However, many immunocompromised patients remain at risk of serious outcomes from COVID-19.

It was established early in the pandemic that immunocompromised COVID-19 patients had higher mortality compared to otherwise healthy COVID-19 patients [1]. Particular risk groups include cancer patients, patients receiving dialysis, and those living with an organ transplant. Several studies have demonstrated that immunocompromised patients exhibit incomplete vaccine response [2] and that they cannot benefit from all available COVID-19 specific treatments, e.g. ritonavir-enhanced nirmatrelvir is associated with significant interactions with many medications used by immunocompromised patients[3].

Here we show that kidney patients, exampled by data from the UK Kidney Association (UKKA), remain at an increased risk for succumbing to COVID-19, compared to the general population in England [4]. This highlights the need for additional protective measures for this population, in particular, as the world begins to lift pandemic-imposed restrictions.

COVID-19 has hit the adult and elderly population disproportionally. The kidney patient population tracked by the UKKA is comprised of adults (≥18), and in 2019 81% of the population was 45 years or older. To minimize age bias, we compared the age distribution for ages 45 and above in both the general population (England) and the kidney patient population (England, Wales and Northern-Ireland) (supplemental table 1). Within the 45 years and older population the estimated average ages are 63.5 for the general population and 64 for the kidney patient population. To have a more conservative approach, we focus our analysis on the general population of ≥45 years and compare them to the adult kidney patient population (≥ 18 years), as data by age group for COVID-19 is not available for kidney patients.

Since the registered COVID-19 deaths are defined as “*an individual who died within 28 days of a positive laboratory result for SARS-CoV-2*”, this number could include deaths that are unrelated to COVID-19, i.e., falsely including patient that died “with” instead of “due to” COVID-19. To account for this, we calculated the expected monthly mortality rate (monthly deaths per 100’000 people) specific for the SARS-CoV-2 positive population by using the reported COVID-19 cases and 2019 mortality rate of both the kidney patient population as well as the general population (herein defined as “monthly background mortality”). This number represents the number of people that would be expected to die in the absence of COVID-19 in the specific population (supplemental table 2).

Next, we analyzed the monthly excess COVID-19 mortality rate in both groups, for each of the three waves (alpha, delta and omicron). We subtracted the calculated monthly background mortality rate from the reported COVID-19 deaths to arrive at the monthly excess COVID-19 mortality rate (excess deaths per 100’000 people) for both populations (Figure 1B).

**Figure 1:**
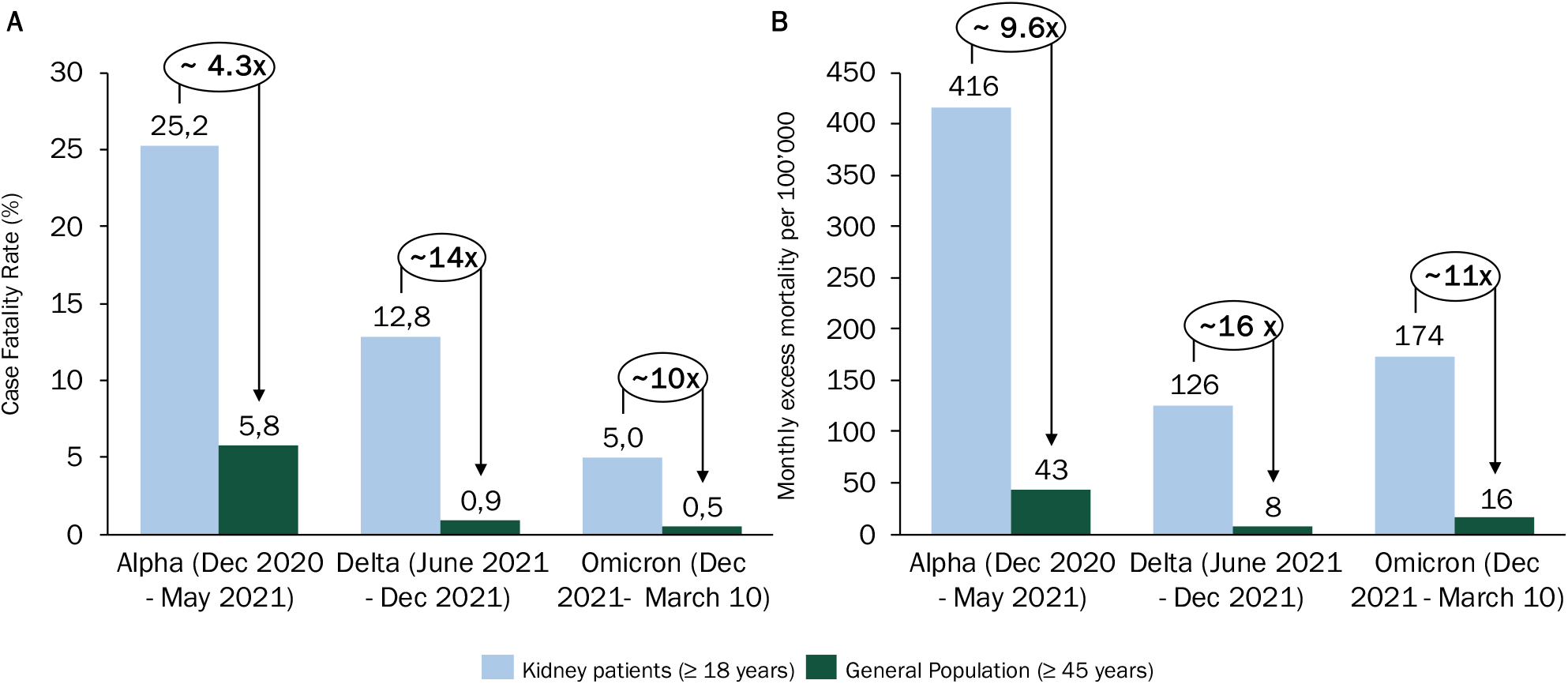
Impact of COVID-19 on Kidney patients and the general population. (A) Case fatality rate and (B) Monthly excess mortality for the three dominant waves in the UK. Analysis under age- and COVID-19 mortality adjustment.

The adjusted monthly mortality rate has consistently been roughly ten-fold higher for the kidney patient population as compared to the general population throughout the pandemic. In the beginning of the pandemic, we found 416 monthly deaths per 100’000 kidney patients more than would have been expected based on the 2019, pre-COVID-19, mortality rate. The monthly excess mortality decreased throughout the pandemic but remained high with 174 monthly excess deaths per 100’000 kidney patients in the omicron wave. The excess mortality during omicron represents a 27% increase in the monthly mortality rate in the kidney patient population relative to pre-COVID19 (2019) observed mortality rate in the same population.

Additionally, we analyzed the case fatality rate as the proportion of excess COVID-19 deaths out of the total COVID-19 reported cases per wave both for kidney patients and general population (Figure 1A).

During the alpha wave as many as 1 in 4 kidney patients who were infected with COVID-19 died, whereas the general population was affected four times less hard (i.e., 1 in 16). Over time we can see decreasing case fatality rates for both groups, though strikingly the case fatality rate in the omicron wave remains ten times higher for kidney patients than the general population.

While our analysis is limited by the lack of patient-level data and additional COVID-19 risk factors we find that kidney patients have a substantially increased risk of dying from COVID-19. We observe a time dependent decline in the case fatality rate, which has decreased from 25.2 to 5.0% for the kidney patient population and from 5.8 to 0.5% for the general population. Due to increased prevalence of COVID-19, mortality rates have not seen a similar decline, indicating that overall, the risk of succumbing to COVID-19 has increased during the recent omicron wave as compared to the delta wave.

The presented results are in striking contrast with the general sentiment of a pandemic under control, where the vaccine and treatment options available suffice and the dominant virus variant has become mild. Many countries have reduced COVID-19 containment measures including reduced access to testing and no mandates on wearing personal protective gear. Under these circumstances it becomes even more important to protect the vulnerable, immunocompromised population from COVID-19. This could be done through special targeted prophylaxis treatment opportunities and adjusted vaccination schemes.

## Data Availability

All data produced in the present study are available upon reasonable request to the authors.

https://ukkidney.org/audit-research/publications-presentations/report/covid-19-surveillance-reports

https://coronavirus.data.gov.uk/

## Disclaimer for UKKA data

The data reported here have been supplied in part by the UKRR of the Renal Association. The interpretation and reporting of these data are the responsibility of the authors and in no way should be seen as an official policy or interpretation of the UKRR or the Renal Association.

## Data availability statement

All data used for this manuscript is publicly available. Analysis can be shared upon reasonable request. The results presented in this paper have not been published previously in whole or part.

