## Supplemental document for "Kidney patients remain at increased risk for succumbing to COVID-19"

**Supplemental Tables**

Table 1: Age distribution in percentage in the kidney patient population compared to the general population of England (≥ 45 years)

| | **Kidney patients** | | **General population England** |
|---|---|---|---|
| **Age range (years)** | **≥ 18 years** | **≥ 45 years** | **≥ 45 years** |
| **18-34** | 7.9 | - | - |
| **35 - 44** | 11.1 | - | - |
| **45 - 54** | 19.5 | 24.1 | 30.1 |
| **55 - 64** | 24.5 | 30.3 | 27.9 |
| **65 - 74** | 21.3 | 26.3 | 22.4 |
| **75 – 84** | 13.1 | 16.1 | 13.9 |
| **≥ 85** | 2.6 | 3.2 | 5.6 |
| **Mean age (calculated estimate)** | 58.3 | 64 | 63.5 |

Table 2: Monthly COVID-19 cases and monthly mortality per 100’000 kidney patients or general population. Monthly mortality rates in 2019 were 649/100’000 in the kidney patient population and 164/100’000 for the general population (≥ 45 years)

| | **Kidney patients – adults (≥ 18 years)** | | | | **General population – adults (≥ 45 years)** | | | | **Fold difference KP vs GP** |
|---|---|---|---|---|---|---|---|---|---|
| **Wave** | **Monthly cases (reported)** | **Monthly COVID-19 deaths (reported)** | **Monthly ‘background’ mortality (calculated)** | **Monthly excess COVID-19 mortality (calculated)** | **Monthly cases (reported)** | **Monthly COVID-19 deaths (reported)** | **Monthly ‘background’ mortality (calculated)** | **Monthly excess COVID-19 mortality (calculated)** | **Adjusted COVID-19 mortality (calculated)** |
| **Alpha** | 1652.98 | 427.13 | 10.73 | 416.39 | 742.94 | 44.60 | 1.22 | 43.38 | 9.6x |
| **Delta** | 985.38 | 132.12 | 6.4 | 125.72 | 916.65 | 9.42 | 1.50 | 7.92 | 16x |
| **Omicron** | 3460.22 | 196.07 | 22.47 | 173.60 | 3100.38 | 19.10 | 2.91 | 16.19 | 11x |

**Supplemental Material & Methods**

For this analysis we relied on publicly available data from England (general population) and the UK Kidney Association (UKKA, kidney patients). Here we refer to kidney patients as (i) patients with chronic kidney disease and (ii) patients on renal replacement therapy, who are under the care of an adult renal center. We used the latest available UKKA Year report (2019) for the mortality rate in the UK kidney and UK general population, and for the age distribution of kidney patients. Data on the age distribution of the general population (England in 2020) was taken from Statista. For weekly numbers on COVID-19 registered deaths and COVID-19 cases we relied on the UKKA COVID-19 surveillance data reports (ukkidney.org) for the kidney patient population, and on the Office for National Statistics (ons.gov.uk) for the general population. We included data until the first week of March 2022. To determine the dominant SARS-CoV-2 variant of concern over time we used GISAID data through covariants.org. We stratified the COVID-19 pandemic by the three major waves to date in the UK; the alpha wave (December 15th 2020 – May 2021), the delta wave (June 2021 – December 10th 2021) and the omicron wave (December 11th 2021 – March 2022). All analyses were performed with Microsoft Excel.
